# Baseline neurological exposome characteristics in a nationwide community-based screening cohort for movement disorders in marginalized and integrated Roma populations

**DOI:** 10.64898/2026.06.30.26356866

**Authors:** Dominika Fricova, Andrej Belak, Jan Necpal, Matus Ferenc, Maria Giertlova, Marek Krauz, Rudolf Balko, Angela Baranova, Martin Buransky, Petra Drencakova, Milan Grofik, Ladislav Gurcik, Vladimir Han, Jozef Haring, Stanislava Jaselska, Bibiana Jelenova, Gabriela Kalafusova, Marek Koren, Kristina Kulcsarova, Alice Kusnirova, Alexandra Lackova, Juraj Mankos, Zuzana Matiskova, Zuzana Mazerikova, Martin Mistrik, Katarina Okalova, Stefan Orkuty, Miriam Ostrozovicova, Jana Papikova, Orest Shabatiuk, Lenka Trckova, Matej Skorvanek, the PURE-MD study group

## Abstract

**Background:** Roma, Europe’s largest ethnic minority, are almost absent from movement-disorder research despite living in environments with biomass combustion, poor sanitation, and unprotected water, creating a high-risk neurological exposome.

**Objective:** To describe baseline exposome profiles in a nationwide Roma screening cohort and compare exposures between marginalized and integrated communities, and between screen-positive and screen-negative participants.

**Methods:** A nationwide community-based program combined door-to-door recruitment of marginalized Roma with recruitment of integrated Roma through neurological services. Participants completed standardized exposome questionnaires and a brief, non-diagnostic motor symptom screen.

**Results:** Among 541 Roma (350 marginalized; 191 integrated), marginalized participants showed greater environmental and socioeconomic disadvantage, and screen-positive individuals tended to cluster at the more adverse end of this exposome spectrum.

**Conclusion:** This nationwide Roma movement-disorder screening cohort with exposome assessment reveals substantial disparities relevant for future clinical and genetic research.

---

Parkinson’s disease (PD) is projected to affect around 20 million individuals worldwide by 2040, with a particularly steep rise in low- and middle-income countries.^1^ Despite this growing burden, clinical cohorts and disease-modifying trials remain strikingly homogeneous: in a meta-analysis of 37 double-blind, randomised, placebo-controlled PD trials, 98% of participants identified as White, while Asian, Black, and Hispanic participants together accounted for less than 2%, and education or rural versus urban residence were rarely reported.^2^ This blind spot is especially relevant for the Roma, Europe’s largest ethnic minority (~10–14 million), who are virtually absent from movement-disorder epidemiology and trials yet often live in segregated settlements marked by poverty, overcrowded and substandard housing, biomass combustion, poor sanitation, unprotected water, unhealthy dietary patterns, and chronic psychosocial stress—features that constitute a distinctive high-risk neurological exposome.^3–5^ Here, we report baseline exposome findings from a nationwide Roma movement-disorder screening cohort in Slovakia (Figure 1), comparing marginalized and integrated communities and contrasting screen-positive with screen-negative participants.

**Figure 1.**
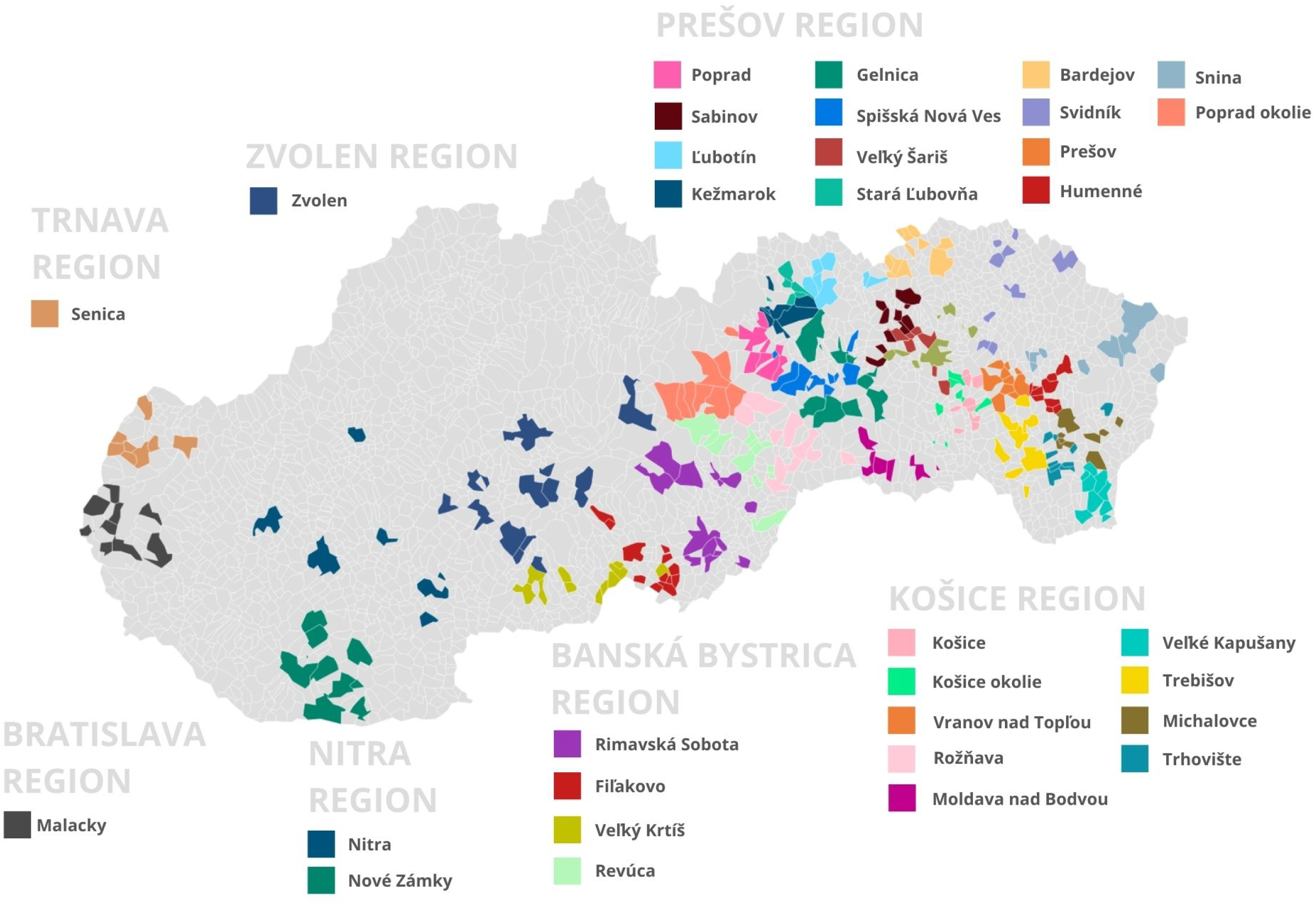
Roma communities covered by national Healthy Regions programme assistants involved in the project

## Methods

### Study design and participants

This ongoing cross-sectional, community-based participatory study was conducted in Slovakia with institutional ethics approval and written informed consent from all participants. Marginalized Roma were recruited through door-to-door screening by trained Roma community health workers within the national Healthy Regions programme, covering more than 450 segregated settlements across all eight administrative regions (Figure 1). Integrated Roma were recruited through neurology outpatient clinics, collaborating neurologists and general practitioners, and patient organisations. For this analysis, we included 350 marginalized and 191 integrated Roma enrolled between January 2023 and 1 February 2026.

### Motor symptom screening and group definitions

In both strata, movement problems were first assessed with the same structured questionnaire, in which participants self-reported slow movements or stiffness, involuntary excessive movements, balance or coordination problems, tremor or jerks, and gait disturbance. These responses are shown in Table 1 as symptom frequencies and reflect self-assessment.

**Table 1.** Demographic, motor, and exposome characteristics of marginalized and integrated Roma participants in PURE-MD.

|  | Characteristic | Marginalized<br>screen-positive<br>participants | Marginalized<br>screen-negative<br>participants | Marginalized<br>total | Integrated<br>screen-positive<br>participants | Integrated<br>screen-negative<br>participants | Integrated<br>total |
| --- | --- | --- | --- | --- | --- | --- | --- |
| DEMOGRAPHICS | N examined | 192 | 158 | 350 | 98 | 93 | 191 |
| | Age, years, mean ± SD | 34.44 ± 22.75 | 44.1 ± 16.85 | 38.82 ± 20.82 | 43.8 ± 23.67 | 42.78 ± 12.94 | 43.31 ± 19.21 |
|  | Female sex, % | 58.85% | 67.09% | 62.57% | 56.12% | 68.82% | 62.30% |
|  | Married, n (%) | 57 (35.19%) | 81 (57.04%) | 138 (45.39%) | 30 (44.12%) | 17 (60.71%) | 47 (48.96%) |
| | No. of children, mean ± SD | 3.59 ± 3.01 | 3.86 ± 0 | 3.73 ± 3.08 | 3.3 ± 2.71 | 2.93 ± 1.8 | 3.17 ± 2.38 |
| | Years of schooling, mean ± SD | 8.4 ± 2.67 | 9.24 ± 2.1 | 8.81 ± 2.44 | 8.54 ± 3.9 | 10.6 ± 4.08 | 9.22 ± 4.05 |
|  | Long-term unemployed, n (%) | 46 (36.51%) | 51 (41.13%) | 97 (38.80%) | 8 (16.00%) | 3 (11.11%) | 11 (14.29%) |
|  | Disability pension, n (%) | 35 (27.78%) | 23 (18.55%) | 58 (23.20%) | 14 (28.00%) | 1 (3.70%) | 15 (19.48%) |
| MOTOR<br>SYMPTOMS<br>SCREENING | Slow movements and stiffness, n (%) | 96 (50.00%) | 67 (42.41%) | 163 (46.57%) | 34 (34.69%) | 0 (0.00%) | 34 (17.80%) |
|  | Involuntary excessive movements, n (%) | 64 (33.33%) | 25 (15.82%) | 89 (25.43%) | 40 (40.82%) | 0 (0.00%) | 40 (20.94%) |
|  | Balance/coordination problems, n (%) | 94 (48.96%) | 65 (41.14%) | 159 (45.43%) | 34 (34.69%) | 0 (0.00%) | 34 (17.80%) |
|  | Tremor or jerks, n (%) | 69 (35.94%) | 36 (22.78%) | 105 (30.00%) | 47 (47.96%) | 0 (0.00%) | 47 (24.61%) |
|  | Gait disturbance, n (%) | 57 (29.69%) | 35 (22.15%) | 92 (26.29%) | 27 (27.55%) | 0 (0.00%) | 27 (14.14%) |
| LIVING<br>ENVIRONMENT | Lived mostly in city, n (%) | 17 (9.34%) | 20 (14.08%) | 37 (11.42%) | 23 (39.66%) | 10 (35.71%) | 33 (38.37%) |
|  | Lived mostly in rural area, n (%) | 49 (26.92%) | 33 (23.24%) | 82 (25.31%) | 25 (43.10%) | 14 (50.00%) | 39 (45.35%) |
|  | Lived in shack/temporary dwelling, n (%) | 31 (17.03%) | 25 (17.61%) | 56 (17.28%) | 0 (0.00%) | 0 (0.00%) | 0 (0.00%) |
| | Household size, mean ± SD | 5.53 ± 3.01 | 5.26 ± 2.9 | 5.41 ± 2.96 | 4.04 ± 2.53 | 4.52 ± 2.26 | 4.2 ± 2.44 |
| | Rooms per household, mean ± SD | 2.62 ± 1.53 | 3 ± 1.51 | 2.79 ± 1.53 | 3.76 ± 2.09 | 3.33 ± 1.14 | 3.62 ± 1.83 |
| WATER | Piped water in dwelling, n (%) | 91 (55.15%) | 82 (63.08%) | 173 (58.64%) | 41 (73.21%) | 21 (75.00%) | 62 (73.81%) |
|  | Own well, n (%) | 23 (13.94%) | 16 (12.31%) | 39 (13.22%) | 11 (19.64%) | 3 (10.71%) | 14 (16.67%) |
|  | Public water source, n (%) | 35 (21.21%) | 24 (18.46%) | 59 (20.00%) | 4 (7.14%) | 4 (14.29%) | 8 (9.52%) |
|  | Spring/stream/other water source, n (%) | 16 (9.70%) | 8 (6.15%) | 24 (8.14%) | 0 (0.00%) | 0 (0.00%) | 0 (0.00%) |
| HEATING,<br>COOKING | Heating - wood-burning stove, n (%) | 146 (80.66%) | 97 (71.32%) | 243 (76.66%) | 22 (39.29%) | 6 (22.22%) | 28 (33.73%) |
|  | Heating - radiators, n (%) | 33 (18.23%) | 24 (17.65%) | 57 (17.98%) | 29 (51.79%) | 19 (70.37%) | 48 (57.83%) |
|  | Cooking - wood stove, n (%) | 102 (58.96%) | 82 (58.99%) | 184 (58.97%) | 13 (23.21%) | 1 (3.57%) | 14 (16.67%) |
|  | Cooking - gas stove, n (%) | 47 (27.17%) | 49 (35.25%) | 96 (30.77%) | 29 (51.79%) | 11 (39.29%) | 40 (47.62%) |
|  | Cooking - electric stove, n (%) | 24 (13.87%) | 8 (5.76%) | 32 (10.26%) | 14 (25.00%) | 16 (57.14%) | 30 (35.71%) |
| SANITATION | Connected to sewage system, n (%) | 94 (51.93%) | 72 (52.17%) | 166 (52.04%) | 44 (86.27%) | 26 (92.86%) | 70 (88.61%) |
|  | Flush toilet in dwelling, n (%) | 106 (59.55%) | 88 (62.86%) | 194 (61.01%) | 56(100.00%) | 28 (100.00%) | 84 (100.00%) |
|  | Bathroom in dwelling, n (%) | 106 (59.55%) | 93 (65.96%) | 199 (62.38%) | 54 (96.43%) | 28 (100.00%) | 82 (97.62%) |
| TOBACCO<br>EXPOSURE | Household daily smoking, n (%) | 118 (67.82%) | 104 (72.73%) | 222 (70.03%) | 31 (55.36%) | 18 (64.29%) | 49 (58.82%) |
|  | Current smokers, n (%) | 72 (37.50%) | 87 (55.06%) | 159 (45.43%) | 24 (24.49%) | 21 (22.58%) | 45 (23.56%) |
| | Cigarettes per day, mean ± SD | 17.54 ± 12.16 | 17.36 ±10.48 | 17.44 ± 11.52 | 12.08 ± 9.16 | 11.52 ± 6.01 | 11.82 ± 7.76 |
| | Years of smoking, mean ± SD | 17.28 ± 17.4 | 18.51 ±13.51 | 17.89 ± 15.54 | 19.75 ± 19.1 | 11.35 ± 9.53 | 16.63 ± 16.68 |
| PHYSICAL ACTIVITY<br>AND STRESS | No regular physical activity, n (%) | 118 (76.62%) | 85 (68.55%) | 203 (73.02%) | 46 (80.70%) | 12 (42.86%) | 58 (68.24%) |
|  | Stress - death of close relative, n (%) | 71 (36.98%) | 75 (47.47%) | 146 (41.71%) | 20 (20.41%) | 10 (10.75%) | 30 (15.71%) |
|  | Stress - illness of close relative, n (%) | 39 (20.31%) | 63 (39.87%) | 102 (29.14%) | 12 (12.24%) | 7 (7.53%) | 19 (9.95%) |
|  | Stress - debt, n (%) | 21 (10.94%) | 28 (17.72%) | 49 (14.00%) | 3 (3.06%) | 2 (2.15%) | 5 (2.62%) |
|  | Household hunger, n (%) | 14 (7.29%) | 14 (8.86%) | 28 (8.00%) | 0 (0.00%) | 0 (0.00%) | 0 (0.00%) |
|  | Stress - housing problems, n (%) | 22 (11.46%) | 18 (11.39%) | 40 (11.43%) | 4 (4.08%) | 4 (4.30%) | 8 (4.19%) |
|  | Stress - fear for children's future, n (%) | 32 (16.67%) | 50 (31.65%) | 82 (23.43%) | 18 (18.37%) | 5 (5.38%) | 23 (12.04%) |
| DIETARY<br>PATTERNS | Dairy products — daily, n (%) | 72 (60.00%) | 86 (56.58%) | 158 (58.14%) | 45 (57.05%) | 18 (72.00%) | 27 (50.00%) |
|  | Processed / smoked meat — daily, n (%) | 82 (62.60%) | 88 (57.89%) | 170 (60.14%) | 24 (32.00%) | 9 (36.00%) | 15 (30.00%) |
|  | Fresh fruit & vegetables — daily, n (%) | 67 (56.30%) | 74 (53.24%) | 141 (54.73%) | 50 (63.29%) | 20 (80.00%) | 30 (55.56%) |
|  | Sweets & sugary drinks — daily, n (%) | 57 (48.31%) | 73 (54.01%) | 130 (51.37%) | 33 (43.42%) | 11 (45.83%) | 22 (42.31%) |
|  | Bread / flour products — daily, n (%) | 111 (86.72%) | 135 (84.37%) | 246 (85.42%) | 68 (85.00%) | 21 (80.77%) | 47 (86.79%) |
|  | Dairy products — daily, n (%) | 72 (60.00%) | 86 (56.58%) | 158 (58.14%) | 45 (57.05%) | 18 (72.00%) | 27 (50.00%) |
| ACCESS TO<br>HEALTHCARE | Ever examined by neurologist, n (%) | 48 (71.64%) | 34 (59.65%) | 82 (66.13%) | 61 (95.31%) | 2 (33.33%) | 63 (90.00%) |
|  | Regular neurologist follow-up, n (%) | 23 (33.33%) | 21 (36.84%) | 44 (34.92%) | 55 (83.33%) | 1 (33.33%) | 56 (81.16%) |
|  | Ever examined by clinical geneticist,<br>n (%) | 7 (10.29%) | 2 (3.57%) | 9 (7.26%) | 30 (46.88%) | 0 (0.00%) | 30 (44.12%) |

In marginalized communities, trained field workers then reviewed these complaints and classified participants as screen-positive or screen-negative; in integrated communities, this classification was performed or supervised by neurologists. Screen-positive marginalized participants were labelled MP (marginalized screen-positive) and screen-negative MHC (marginalized healthy controls); the corresponding integrated groups were IP (integrated screen-positive) and IHC (integrated healthy controls), with MT (marginalized total) and IT (integrated total) denoting the full marginalized and integrated samples.

Because classification was based on this structured review, some screen-negative individuals (especially in the marginalized group) still report motor-like difficulties that were judged more consistent with non-neurological, mainly musculoskeletal problems. Motor data presented here are therefore screening-level findings and do not yet represent neurologist-confirmed diagnoses.

### Exposome assessment

All participants completed a standardised questionnaire covering exposome domains with direct neurological relevance. Collected data included demographic and socioeconomic characteristics (age, sex, marital status, number of children, years of schooling, long-term unemployment, disability pension), living environment and housing (urban vs rural residence, dwelling type, household size, number of rooms), drinking water sources (household tap, private well, public community source, spring or other unprotected source), indoor combustion (heating and cooking fuels, including wood, gas, electricity, and radiators/central heating), sanitation infrastructure (connection to sewage system, presence of flush toilet and bathroom), tobacco exposure (household daily smoking, current smoking, cigarettes per day, years of smoking), dietary patterns (daily intake of dairy products, processed/smoked meat, fresh fruit and vegetables, sweets/sugary drinks, and bread/flour products), physical activity, and psychosocial stressors (death or illness of close relatives, debt, housing problems, household hunger, fear for children’s future). Access to neurological and genetic healthcare, family history of neurological disease, and parental consanguinity were also recorded. Data were captured in a secure, GDPR-compliant REDCap database.

### Descriptive analyses

Continuous variables are summarized as mean ± standard deviation and categorical variables as number (percentage). The present report focuses on describing the distribution of demographic, motor, and exposome characteristics between marginalized and integrated participants and between screen-positive and screen-negative groups within each stratum.

## Results

### Cohort characteristics

By 1 February 2026, 541 Roma had been enrolled, including 350 marginalized participants (MT) and 191 integrated participants (IT). As summarized in Table 1, marginalized participants were generally younger, more often long-term unemployed, and had slightly fewer years of schooling than integrated participants. Within the marginalized stratum, 192 participants were classified as screen-positive (MP) and 158 as screen-negative (MHC), while within the integrated stratum, 98 were screen-positive (IP) and 93 screen-negative (IHC) (Table 1).

### Exposome differences: marginalized versus integrated

Across most domains, MT had a more adverse exposome profile than IT, including larger and more crowded households and less frequent access to piped water and sewage connections, while temporary or shack housing and the use of unprotected springs or streams as water sources occured exclusively in marginalized settlements. Indoor combustion showed the clearest gradient: wood-burning remained the dominant heating and cooking fuel in MT, whereas IT more often used radiators and electric or gas stoves; tobacco exposure was also higher in MT, with more current smokers, greater household daily smoking, and higher cumulative tobacco use. Physical inactivity and several psychosocial stressors, including household hunger and debt-related or housing-related stress, were also more frequent in MT, indicating a denser clustering of environmental and social disadvantages. Access to specialist care followed the opposite pattern, with IT more often reporting prior neurological assessment, regular neurologist follow-up, and examination by a clinical geneticist than MT (Table 1).

### Exposome differences by screening status

Within the marginalized stratum, MP showed a somewhat more adverse profile than MHC despite living in the same settlements, including younger age, lower educational attainment, higher disability pension rates, fewer rooms per household, greater reliance on wood-burning heating, and less regular physical activity (Table 1). Overall, these patterns suggest a modest clustering of social and environmental disadvantage among screen-positive participants within marginalized communities.

Within the integrated stratum, contrasts between IP and IHC were more pronounced. IP had lower educational attainment, higher disability pension rates, greater reliance on wood for heating and cooking, less frequent use of electric stoves, and markedly lower levels of regular physical activity than IHC (Table 1). Together, these findings indicate that screen-positive participants in both strata tended to occupy the less favorable end of the exposome spectrum.

### Motor symptom screening

By design, screen-positive groups had higher self-reported motor symptom burden than screen-negative groups, but the pattern of specific complaints differed between strata. Among MT, both MP and MHC frequently reported slow movements and stiffness, balance or coordination problems, tremor, and gait disturbance, although symptom frequencies were consistently higher in MP; as described in Methods, many complaints in MHC were considered more consistent with pain-related or orthopaedic problems and are therefore best interpreted as screening-level motor symptoms rather than confirmed movement-disorder signs.

In IT, the most frequent motor complaints in IP were tremor, involuntary excessive movements, and slow movements and stiffness, whereas, by screening definition, IHC did not report these key motor symptoms (Table 1). All motor findings presented here are based on self-reported screening questionnaires and will be re-evaluated with full neurological examinations in subsequent study phases.

## Discussion

This brief report provides the first systematic description of a neurological exposome in Roma communities undergoing nationwide movement-disorder screening. Marginalized Roma were consistently exposed to a dense cluster of environmental and social risk factors, including biomass combustion, inadequate water and sanitation, high tobacco exposure, unfavourable dietary patterns, and chronic psychosocial stress, mirroring broader evidence of structural disadvantage and health inequities in Slovak Roma settlements.^4,5^ These findings suggest that Europe’s largest ethnic minority is not only underrepresented in movement-disorder research but also embedded in environments that may heighten susceptibility to neurodegeneration.^1,2,6^

Our data indicate that screen-positive individuals in both strata tend to cluster at the most adverse end of the exposome spectrum, with younger age, lower education, higher disability rates, more crowded living conditions, and less healthy diets than screen-negative participants in the same communities. Within both strata, screen-positive participants more often reported daily consumption of processed or smoked meat and lower intake of fresh fruit and vegetables, raising the hypothesis that cumulative environmental disadvantage contributes to early movement-related complaints, even though screening status alone does not represent a neurological diagnosis.

Genetic susceptibility is likely to intersect with this exposome. Roma populations are known to harbour founder mutations in genes linked to movement disorders and other neurological conditions, reflecting historical isolation and episodes of consanguinity.^3,7–9^ The elevated rates of parental consanguinity observed in marginalized Roma, combined with concentrated environmental risk in segregated settlements, create conditions in which recessive variants and environmental insults may co-occur, complicating attempts to distinguish genetic from environmental drivers of disease. Future work will therefore integrate standardised neurological examinations, detailed symptom phenotyping, and genomic analyses to explore gene–environment interactions underlying movement disorders in these communities.^3,7–10^

Limitations of this study include its cross-sectional design and reliance on self-reported exposures and a brief screening tool rather than full neurological examination. Screening categories were based on self-reported motor complaints reviewed by trained staff, and screen-negative groups— especially in marginalized settlements—include individuals whose motor-like difficulties were judged more consistent with musculoskeletal or other non-neurological problems, so “motor symptoms” in these participants represent a heterogeneous screening-level signal rather than a definitive movement-disorder phenotype. Nonetheless, the present baseline provides an exposome framework for future longitudinal analyses of movement disorders, including comparisons of clinical phenotypes between marginalized and integrated Roma and between screen-positive and screen-negative groups.

In summary, this study documents substantial environmental and social disparities between marginalized and integrated Roma and suggests that individuals screening positive for movement symptoms, particularly in integrated settings, cluster at the high-risk end of this exposome gradient. Linking detailed exposure profiles with subsequent clinical and genetic evaluation will help clarify how neurological disease in Roma populations emerges from the interplay of environmental adversity, social structure, and inherited risk, and can bring movement-disorder research closer to the diversity of the populations it aims to serve.^1–3,7–10^

## Data Availability

Anonymised data will be made available upon reasonable request following completion of primary PURE-MD analyses.

## References

1. Su D, et al. Projections for prevalence of Parkinson’s disease and its driving factors in 195 countries and territories to 2050: modelling study of Global Burden of Disease Study 2021. BMJ. 2025;388:e080952.

2. Rodrigues FR, Bolner G, Ribeiro GBS, et al. Racial and ethnic diversity in clinical trials for disease-modifying drugs in Parkinson disease: a systematic review and meta-analysis. Mov Disord Clin Pract. 2026;13(5):1295–1300.

3. Kalaydjieva L, Gresham D, Calafell F. Genetic studies of the Roma (Gypsies): a review. BMC Med Genet. 2001;2:5.

4. Bosakova L, et al. Association of socioeconomic disadvantage and ethnicity with perinatal, neonatal, and infant mortality in Slovakia. Int J Equity Health. 2024;23:85.

5. Belak A, et al. Why don’t health care frontline professionals do more for segregated Roma? Soc Sci Med. 2020;246:112739.

6. Aamodt WW, Willis AW, Dahodwala N. Racial and ethnic disparities in Parkinson disease. Neurol Clin Pract. 2023;13(2):e200138.

7. Ostrozovicova M, et al. A recurrent VPS16 p.Arg187* nonsense variant in early-onset generalized dystonia. Mov Disord. 2021;36:1984–1985.

8. Hamilton EMC, et al. UFM1 founder mutation in the Roma population causes a recessive variant of H-ABC. Neurology. 2017;89(18):1821–1828.

9. Olgiati S, et al. DNAJC6 mutations associated with early-onset Parkinson’s disease. Ann Neurol. 2016;79(2):244–256.

10. Schumacher-Schuh AF, et al. Underrepresented populations in Parkinson’s genetics research. Mov Disord. 2022;37(8):1593–1604.

